# Supra-second interval timing deficits and abnormal frontal theta oscillations in individuals with bipolar disorder

**DOI:** 10.1101/2022.09.25.22280348

**Authors:** Victόria A. Müller Ewald, Nicholas T. Trapp, McCall E. Sarrett, Benjamin D. Pace, Jenny G. Richards, Ilisa K. Gala, Jacob N. Miller, Jan R. Wessel, Vincent A. Magnotta, John A. Wemmie, Aaron D. Boes, Krystal L. Parker

## Abstract

**Objectives:** Though widely reported by patients, cognitive symptoms associated with bipolar disorder (BD), including deficits in executive function, memory, attention, and timing are under-studied. Work suggests that individuals with BD show impairments in sub-second interval timing tasks (ITT), however, results have been inconclusive regarding supra-second time perception in BD patients. Additionally, the effects of mood or medication status on time perception in BD patients are debated in the literature.

**Methods:** To address this, the present work administered a supra-second ITT concurrent with electroencephalographic (EEG) recordings to patients with BD and neuronormative controls. As this task is known to elicit frontal theta oscillations, which can be abnormal in psychiatric populations, signal from the Fz lead was analyzed at rest and during the task.

**Results & Conclusions:** As hypothesized, results suggest that individuals with BD show impairments in supra-second ITT performance compared to neuronormative controls. Frontal theta power was also reduced compared to controls during the ITT but not during rest. Finally, timing impairments remain detectible in BD patients independent of mood state and use of antipsychotic medication. This suggests that supra-second interval timing deficits are a key characteristic observed in BD. Together with previous work, these findings point to critical timing impairments in BD patients across a wide range of timing modalities and durations.

## Introduction

The diagnosis and treatment for bipolar disorder (BD) focuses largely on mood symptoms^1-4^. However, changes in cognitive functioning, including deficits in prospective memory, working memory, executive function, attention, planning, and timing^5-7^, are common and may even precede a formal BD diagnosis^8^. Cognitive symptoms are reported by both patients with bipolar I disorder (BDI) and bipolar II disorder (BDII), and are present even when individuals are in a euthymic state^9^. Thus, it is imperative that we develop a better understanding of the cognitive symptoms associated with BD, as they are both linked to a lower quality of life^10^ and widely reported by patients.

Reports suggest that some cognitive impairments in BD can recede with the treatment of mood symptoms^11^. However, other cognitive symptoms, such as impairments in verbal memory and sustained attention, persist or worsen following mood symptom treatment^11,12^. It has been proposed that impairments on measures of general intellectual functioning, working memory, and cognitive set-shifting are associated with antipsychotic medication use in BD patients^6^. These deficits may be attributable to anti-psychotic induced reductions in information processing speed. Understanding how time processing is altered in BD patients, especially as a function of mood or medication status, would be directly informative to this debate.

Previous work suggests that individuals with BD show impaired performance in sub-second interval timing tasks (ITT)^13,14^. Additionally, individuals with BD show abnormal acquisition of single-cue delay eye blink conditioning, indicating deficits in implicit motor timing^15^. Interval timing is dependent on activity in diffuse neural networks including the cerebello-thalamo-cortical network and the cortico-striatal network^16,17^. Work also indicates that abnormalities in nodes of these networks which are necessary for interval timing are involved in the pathophysiology of BD. Such areas include the frontal cortex, thalamus, and cerebellum^18-22^. Despite well-characterized abnormalities in sub-second timing, it is unclear if individuals with BD also present deficits in supra-second timing. Two studies have explored this question, suggesting impairments compared to neuronormative controls^23,24^. However, the population in these studies included individuals with schizophrenia (SCZ) as well as individuals with depression and BD, confounding the interpretation of timing deficits in BD specifically.

Cognitive impairments in SCZ are well characterized and include impairments in executive function, timing, and verbal memory^4^. Previous work from our laboratory suggests that impairments in supra-second interval timing correlate with abnormal fronto-central theta oscillations in patients with SCZ^25,26^. Work suggest significant genetic and symptomatic overlap between SCZ and BD^4,12^. For example, BD patients suffering from mania commonly present with Schneiderian first-rank symptoms, including delusions and hallucinations. Additionally, patients with SCZ may also present with depression or mania^4^. However, it is unclear if the deficits in timing and frontal oscillations observed in SCZ will also extend to BD.

To address these questions, we administered a supra-second ITT to participants with BD and neuronormative controls (CT) while simultaneously recording electroencephalographic (EEG) activity. We hypothesized that individuals with BD would show impaired supra-second ITT performance and reduced frontal theta power compared to the CT group. Participants with BD I and BD II were included in the present study to characterize possible differences between these two populations. Given the similar cognitive profiles between the two patient populations, we did not expect to find differences in ITT performance between the BD I and BD II groups^9^. Finally, we assessed differences in ITT performance and frontal theta in BD depending on mood or medication status.

## Methods

### Subjects

Twenty-four participants (20 females, 4 males) with a DSM-IV diagnosis of BDI (16 subjects) or BDII (8 subjects) were recruited from the Iowa Longitudinal Database (see **Table 1** for demographic information and **Table 2** for comorbidities). All subjects had diagnoses confirmed by a board-certified psychiatrist at the University of Iowa Hospitals & Clinics. Medication status was stable for a minimum of 30 days prior to enrollment and was not altered for the present study (**Table 3**). Individuals who reported illicit drug use within 6 months of study commencement were excluded from participation. Mood was assessed with the Montgomery-Asberg depression rating scale and a depressed state was defined as a score greater than 6^27^. Six CT subjects were included as a neuronormative comparison group. CT subjects did not have a history of neuropsychiatric disorders. In accordance with federal and institutional guidelines, all procedures including informed consent were approved by the University of Iowa Institutional Review Board and are in accordance with the Declaration of Helsinki.

**TABLE 1.** Participant Demographics

| Baseline Characteristic | Controls<br>(n=6) | Bipolar type I<br>(n=16) | Bipolar type II<br>(n=8) | p-value |
| --- | --- | --- | --- | --- |
| Age |  |  |  | 0.948 |
| Mean (SD) | 37.2 (10.3) | 37.2 (13.5) | 35.5 (12.1) |  |
| Sex |  |  |  | 0.212 |
| Female | 3 (50%) | 13 (81.25%) | 7 (12.50%) |  |
| Male | 3 (50%) | 3 (18.75%) | 1 (87.50%) |  |
| Race |  |  |  | 0.471 |
| White | 4 (66.67%) | 12 (75%) | 6 (75%) |  |
| Black/African American | 0 (0%) | 3 (18.75%) | 0 (0%) |  |
| Asian | 1 (16.67%) | 0 (0%) | 1 (12.50%) |  |
| Other | 1 (16.67%) | 1 (6.25%) | 1 (12.50%) |  |
| Education |  |  |  | 0.138 |
| No High School | 0 (0%) | 2 (12.50%) | 0 (0%) |  |
| High School | 0 (0%) | 5 (31.25%) | 3 (37.50%) |  |
| Associate's/Bachelor's | 3 (50%) | 7 (43.75%) | 5 (62.50%) |  |
| Post-Graduate | 3 (50%) | 2 (12.50%) | 0 (0%) |  |
| Handedness |  |  |  | 0.126 |
| Right | 5 (83.33%) | 16 (100%) | 8 (100%) |  |
| Left | 1 (16.67%) | 0 (0%) | 0 (0%) |  |

**TABLE 2.** Comorbidities reported at enrollment

| Comorbidity | N (%) |
| --- | --- |
| Generalized Anxiety Disorder | 5 (20.8%) |
| Post-Traumatic Stress Disorder | 4 (16.67%) |
| Panic Disorder | 2 (8.33%) |
| Borderline Personality Disorder | 2 (8.33%) |
| Attention-Deficit/Hyperactivity Disorder | 2 (8.33%) |
| Migraine | 1 (4.17%) |
| Fibromyalgia | 1 (4.17%) |
| Other Medical Condition | 5 (20.8%) |

**TABLE 3.**
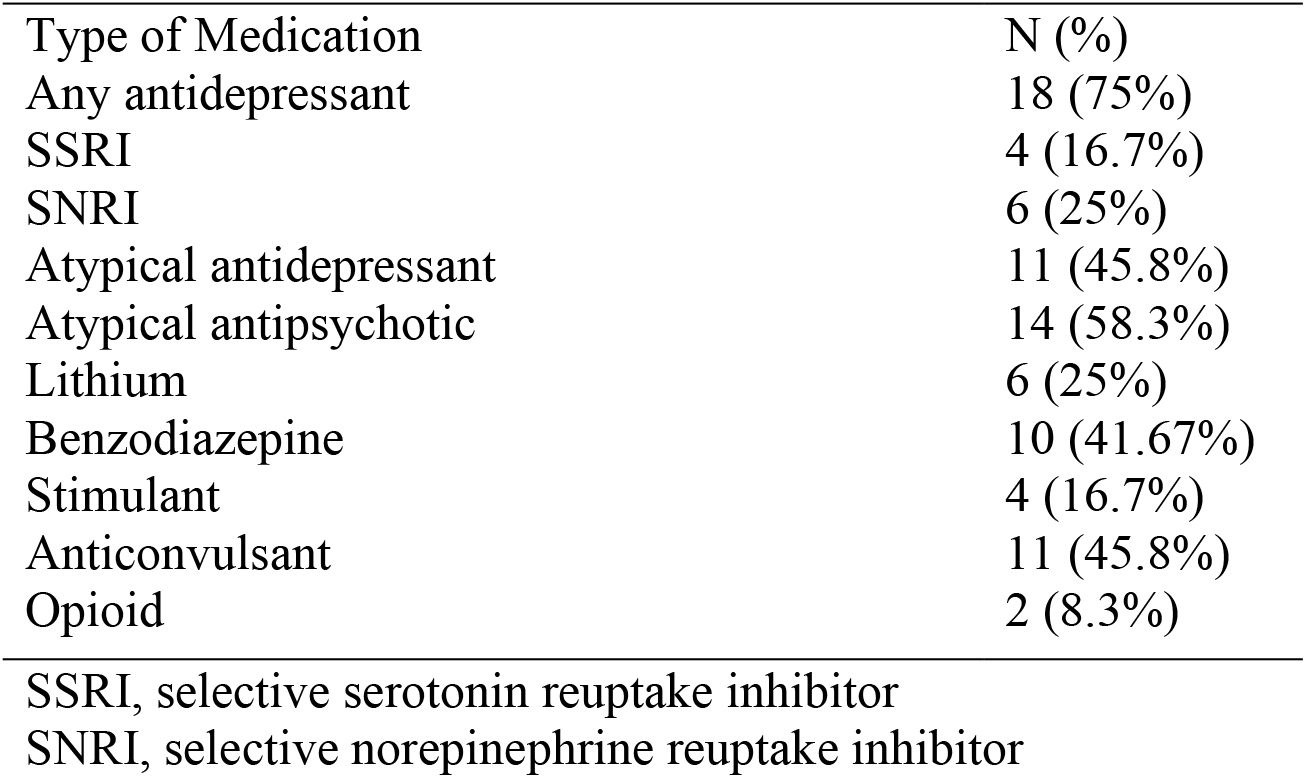
Medications at time of enrollment

### Tasks

#### Interval timing task

Concurrent with EEG acquisition, participants performed a supra-second ITT (**Figure 1A**). Participants completed the task sitting in front of a Dell 20” monitor with a 60 Hz refresh rate. White times new roman size 40 text appeared on a black background in the middle of the screen. Participants received verbal instructions on how to perform the task from the experimenter and read the same set of instructions on the computer screen. As is customary for this task, all participants were instructed not to count time in their head. To start each trial, a number which indicated the interval to be estimated by the participant (“3” for the short interval/SIT or “12” for the long interval/LIT) appeared on the screen. Participants pressed the space bar on the task computer to start the trial and to indicate their judgement of the elapsed interval thus ending the trial. Feedback about response accuracy was shown for every trial immediately following the button press. Feedback consisted of the following information: “you were x.xxx seconds early/late.” Participants pressed the space bar to initiate the next trial. The experiment consisted of a total of 80 trials (40 SIT trials & 40 LIT trials) presented in pseudo-random order.

**Figure 1.**
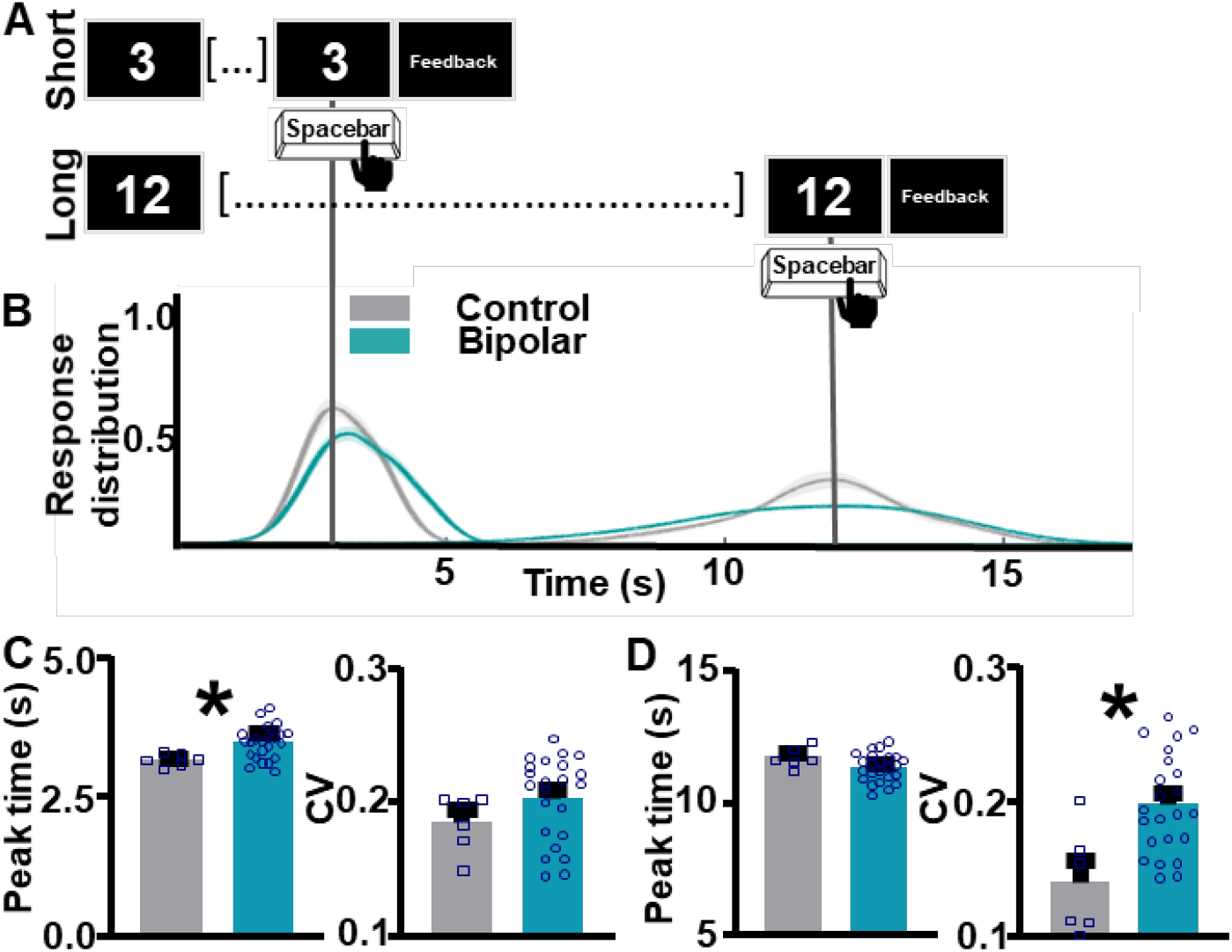
Individuals with bipolar disorder show impairments in supra-second interval timing. **A**. Schematic diagram of supra-second interval timing task. Trials begin when participants are shown a 3s or a 12s timing cue. Participants press the spacebar to indicate their estimation of the target interval. **B**. Response distribution for neuronormative controls vs. individuals with bipolar disorder. **C**. Individuals with bipolar disorder over-estimate the short interval duration compared to controls **[left]**. No differences in response distribution were detected **[right]. D**. Individuals with bipolar disorder do not differ from controls in estimation of the long interval duration **[left]**, however, individuals with bipolar disorder have a significantly wider response distribution compared to controls **[right]**. Mean and standard error of the mean plotted in bar graphs. Dots represent values from individual subjects. * p < 0.05

#### Resting state task

Resting-state recordings lasted 5 minutes, during which participants sat in a chair, were instructed to keep their eyes open, look forward, and to let their mind wander.

### EEG acquisition

A BrainVision 64-channel active electrode system with Ag/AgCl electrodes was used to collect EEG (Morrisville, NC). A custom-made electrode cap was utilized, which included electrode placements that are not typical of the International 10-20 system^28^. Specifically, electrodes PO3 and PO4 were substituted by electrodes I1 and I2 which flanked the Iz electrode. These were situated at the back of the head over the inion, below O1, Oz, and O2. At the beginning of the recordings, impedances were reduced using high viscosity electrode gel for active electrodes (EASYCAP, Munich, Germany). Impedance for all electrodes was kept at or below 15 kΩ for the duration of the recording. Data were acquired at 500 Hz and referenced online to Pz.

### EEG analyses

More detailed analysis information can be found in the sub-sections below. In brief, analyses focused on electrode Fz, as frontal theta oscillations are typically maximal at this electrode^29^. Frequency bands were defined as follows: delta (1-4 Hz), theta (4-8 Hz), alpha (8-13 Hz), beta (13-30 Hz), and gamma (30-50 Hz). For exploratory analyses, the frequency-space assessed was between 1 and 50Hz. The time-window assessed for the short interval was from cue-onset to 3 s post-cue. The time-window assessed for the long interval was from cue-onset to 12 s post-cue. For region of interest (ROI)-based analyses the frequency space assessed was from 4 to 8 Hz (theta band) and the time-window analyzed was the 500 ms following the timing-cue onset.

#### Preprocessing

Data were preprocessed using custom MATLAB scripts based on EEGLAB^30^ functions. Data were high-pass filtered at 1Hz, then low-pass filtered at 50Hz, the transition bandwidth was set to twice the cutoff frequency (−6 dB) for cutoff <= 1Hz and 25% cutoff frequency for cutoff > 8 Hz. Nonstereotypic artifacts were removed manually, which resulted in the exclusion of 18% of trials on average. Next, continuous data were rereferenced offline to the average voltage. Eyeblinks and saccades were removed using independent component analysis. Finally, data were epoched as follows: for short interval cue presentation, data were epoched from 1 second before cue presentation to 5 seconds after cue presentation. For long interval cue presentation, data were epoched from 1 second before cue presentation to 15 seconds after cue presentation. For responses, data were epoched from 1 second before the response to 1 second after the response.

#### Non time-locked analyses

To assess high-level differences between BD and CT groups during the task and at rest, non-time-locked analyses were conducted using the fast-Fourier transform method. Relative power at each frequency band was defined as the proportion of the overall spectral power distribution area occupied by that frequency band. This was quantified using the MATLAB function trapz. For ITT task analyses, short and long interval trials were epoched as described above and analyzed together. For resting-state analyses, continuous data were epoched into 20s intervals.

#### Time-locked analyses

Time-locked analyses were conducted on ITT task data via Morlet wavelet convolution to assess oscillatory activity during specific task epochs. In brief, the power spectrum for each trial was convolved with complex Morlet wavelets with a width of 7 cycles, one wavelet for each frequency (1 to 50 Hz). Power values were baseline normalized by conversion to the decibel (dB) scale (10*log10(power epoch/power baseline)). The 250 ms window from -400 to -150 ms prior to the presentation of the timing cue was utilized as the baseline for both cue and response-centered analyses.

### Statistical analyses

For ITT performance analyses, participants’ time estimates for the SIT/LIT intervals were fit with Gaussian distributions using custom-written MATLAB routines (MathWorks, Natick, MA). Timing accuracy and precision were estimated by calculating peak time and CV measures for each participant, respectively. The peak time index represents the accuracy of participants’ responses and was calculated using the best fit estimate of the Gaussian distribution. The CV index represents the precision of participants’ responses and was calculated by dividing the response standard deviation by peak time.

Exploratory analyses of neural oscillations were conducted in MATLAB using t-tests. A simple false-discovery rate correction was used for cell-by-cell comparison of spectral activity over time. ROI-based analyses of neural oscillations and interval timing task performance were conducted in GraphPad Prism (San Diego, California). T-tests were used for comparisons across two groups. One-way ANOVAs were used for comparisons across three groups. Multiple comparisons were corrected for using Tukey’s multiple comparisons test. Statistical outliers were defined as individuals with scores 2 standard deviations above/below their group mean and excluded from the analysis. For analyses where outliers were excluded, the number of individuals excluded and which group they belonged to are noted in the results section.

## Results

As individuals with BDI and BDII did not differ in ITT performance or frontal theta power (**Supplemental Figure 1**), groups were combined for the main analyses below.

### Interval timing performance differences between CT and BD groups

During the supra-second ITT (**Figure 1A**) individuals with BD showed impaired performance compared to the CT group (**Figure 1B**). For the short interval, individuals with BD show an over-estimation of the target duration compared to the CT group, as quantified by the peak time index (*t*_(27)_ = 2.61, *p* = 0.0146, one BD outlier excluded, **Figure 1C [left]**). Response distribution, quantified by the CV index, does not differ between groups (*t*_(27)_ = 1.338, *p* = 0.1920, one BD outlier excluded, **Figure 1C [right]**). For the long interval, peak response times do not differ between BD and CT groups (*t*_(28)_ = 1.576, *p* = 0.1262, **Figure 1D[left]**). However, individuals with BD showed significantly lower CV indices, indicating higher variability in response times compared to the CT group (*t*_(27)_ = 3.345, *p* = 0.0024, one BD outlier excluded, **Figure 1D[right]**).

### Frontal theta differences between CT and BD groups

Whole head EEG recordings were acquired with data analysis focusing on the Fz electrode (**Figure 2A, red**).

**Figure 2.**
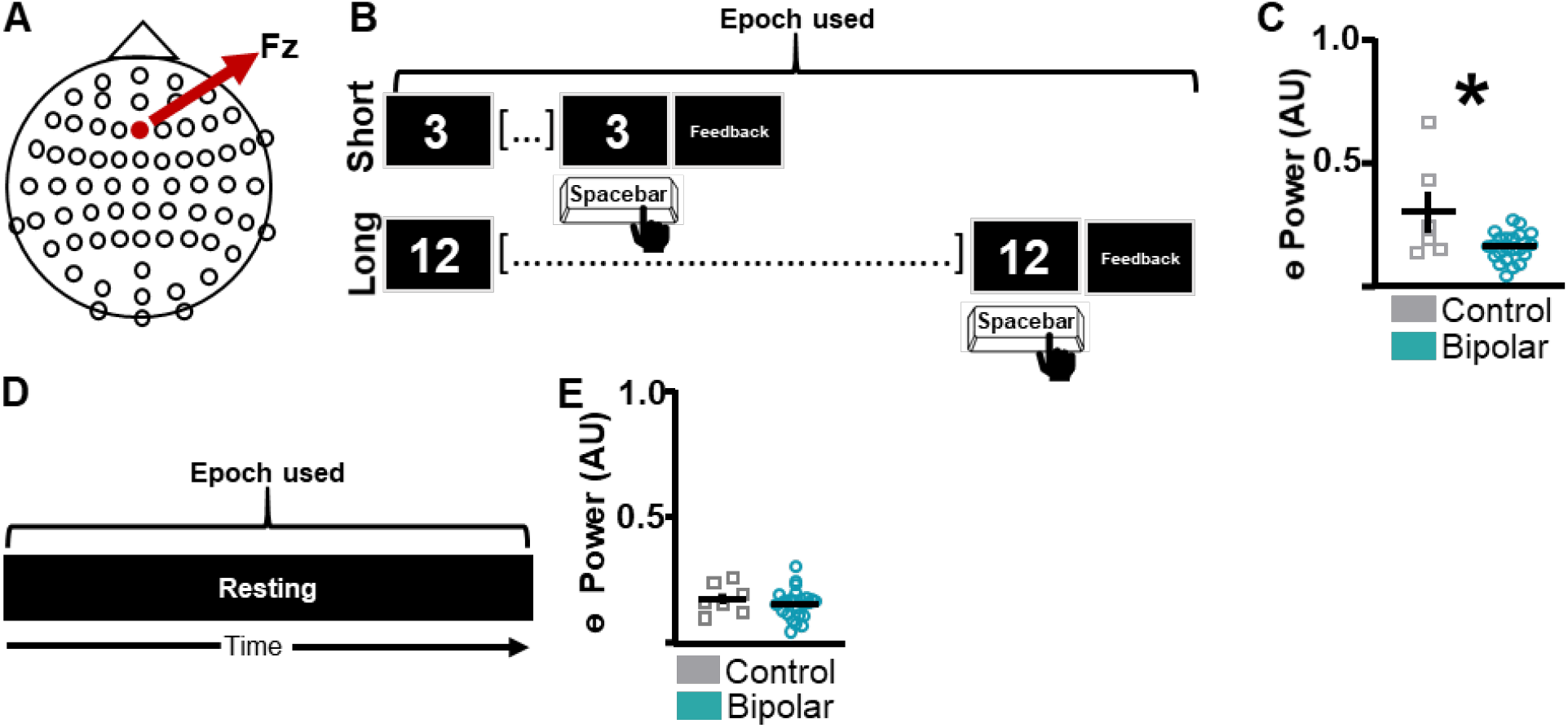
Abnormal frontal theta power in individuals with bipolar disorder compared to neuronormative controls. **A**. Schematic diagram of electrode cap used for data acquisition, with lead of interest (Fz) marked in red. **B**. To assess task-wide differences in oscillatory activity between bipolar disorder and neuronormative control groups data from the whole interval-timing task were analyzed. **C**. Individuals with bipolar disorder show lower theta power compared to individuals in the neuronormative control group during the supra-second interval timing task. **D**. To assess resting-state differences in oscillatory activity between bipolar disorder and neuronormative control groups data from the whole resting-state task were analyzed. **E**. No differences in resting-state theta power were identified between neuronormative control and bipolar groups. Mean and standard error of the mean plotted in bar graphs. Dots represent values from individual subjects. * p < 0.05

#### Task-wide differences – ITT

To assess task-wide differences in oscillatory activity between BD and CT groups, spectral activity during all trial epochs (SIT + LIT) was quantified (**Figure 2B**). During the ITT, individuals with BD showed lower frontal theta power compared to the CT group (*t*_(27)_ = 2.992, *p* = 0.0059, one BD outlier excluded, **Figure 2C**). Although it can be observed that the theta power values of a single CT subject are higher than the remainder of the CT subjects, data from this individual were not excluded as they do not fit the statistical outlier criteria as described in the methods section. Differences in power during the ITT were not observed in other frequency bands (**Supplemental Figure 2**).

#### Task-wide differences – Resting state

To assess if differences in theta power between BD and CT groups were diffuse or task-specific, resting-state data were analyzed (**Figure 2D**). There were no significant differences in theta power between BD and CT groups (*t*_(30)_ = 0.8343, *p* = 0.4107, one CT & one BD outlier excluded, **Figure 2E**) during rest.

Because our group has shown that theta power immediately following timing-cue onset correlates to ITT performance^25^, we next examined time-locked activity.

#### Time-locked analyses – Short interval

Data were epoched surrounding the presentation of the short interval timing cue (**Figure 3A**). Averaged spectrograms were created for individuals in the CT group (**Figure 3B [left]**) and the BD group (**Figure 3B [right]**). Exploratory analyses suggest that oscillatory activity does not differ between groups. ROI-based analyses suggest that post-cue theta power did not significantly differ between groups (*t*_(24)_ = 1.911, *p* = 0.0680, one BD outlier excluded, **Figure 3C**).

**Figure 3.**
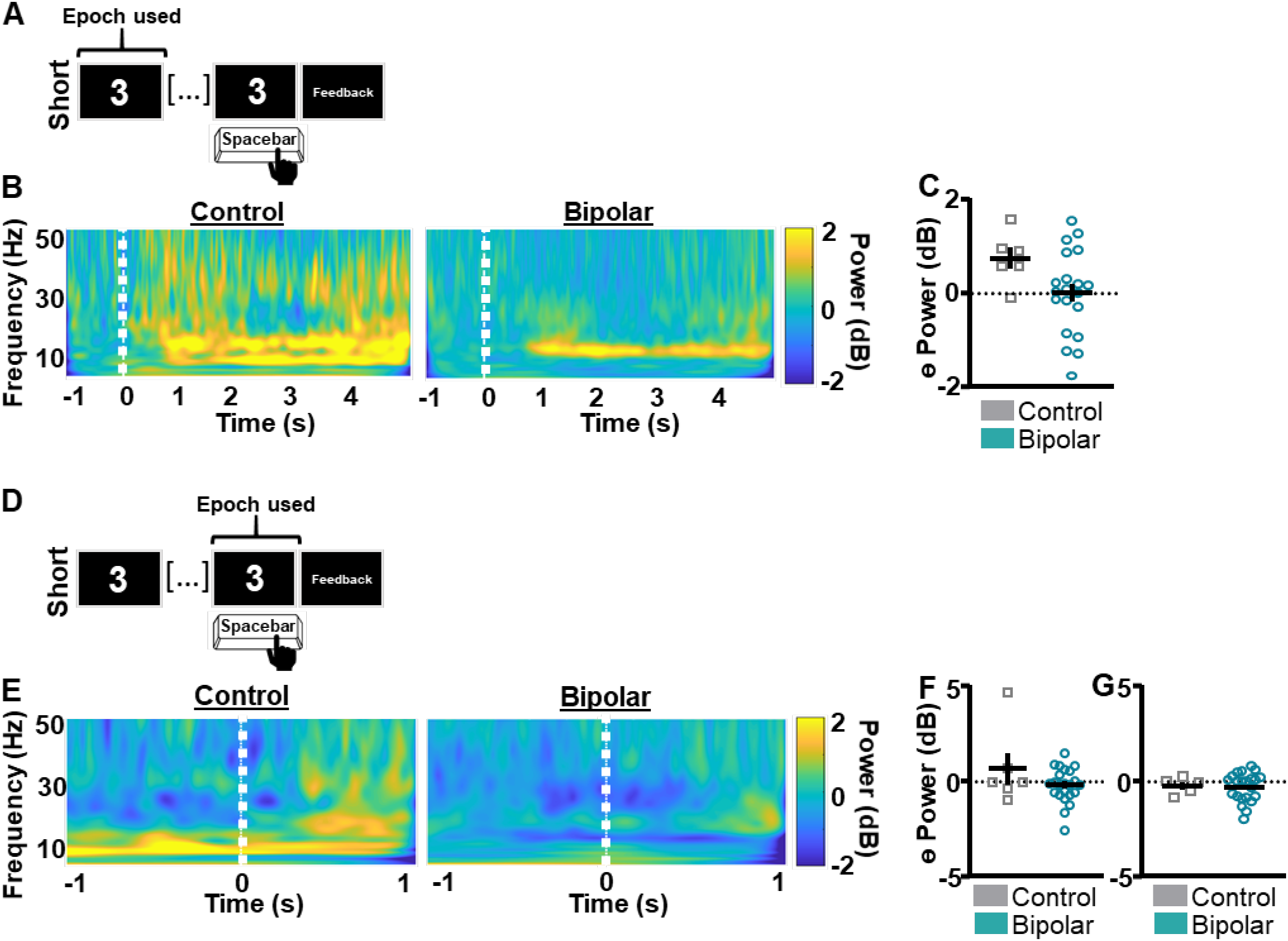
Short interval oscillatory activity does not differ between bipolar and control groups. **A**. Data were epoched around the presentation of the short interval timing cue. **B**. Averaged spectrogram of individuals in the control group **[left]** and the bipolar group **[right]**. Exploratory analyses suggest that oscillatory activity does not differ between the two groups during the whole short interval epoch. **C**. ROI-based analyses indicate that theta power following the timing cue does not differ between bipolar and control groups. **D**. Data were epoched around the short interval button press. **E**. Averaged spectrogram of individuals in the control group **[left]** and the bipolar group **[right]**. Exploratory analyses suggest that oscillatory activity does not differ between the two groups. **F**. ROI-based analyses indicate that theta power prior to the response does not differ between bipolar and control groups. **G**. ROI-based analyses indicate that theta power following the response does not differ between bipolar and control groups. Mean and standard error of the mean plotted in bar graphs. Dots represent values from individual subjects.

Data were next epoched surrounding the button press indicating the participants’ estimation of short interval duration (**Figure 3D**). Averaged spectrograms were created for individuals in the CT group (**Figure 3E[left]**) and the BD group (**Figure 3E[right]**). Exploratory analyses suggest that oscillatory activity did not differ between groups. ROI-based analyses suggest that neither pre-response nor post-response theta power differed between groups (*t*_(27)_ = 1.556, *p* = 0.1313, one BD outlier excluded, **Figure 3F**; *t*_(25)_ = 0.2206, *p* = 0.8272 one CT outlier and & BD outliers excluded, **Figures 3G**).

#### Time-locked analyses – Long interval

Data were epoched surrounding the presentation of the long interval timing cue (**Figure 4A**). Averaged spectrograms were created for individuals in the CT group (**Figure 4B [left]**) and the BD group (**Figure 4B [right]**). Exploratory analyses suggest that oscillatory activity did not differ between groups. ROI-based analyses suggest that post-cue theta power did not differ between groups (*t*_(24)_ = 1.663, *p* = 0.1094, one BD outlier excluded, **Figure 4C**).

**Figure 4.**
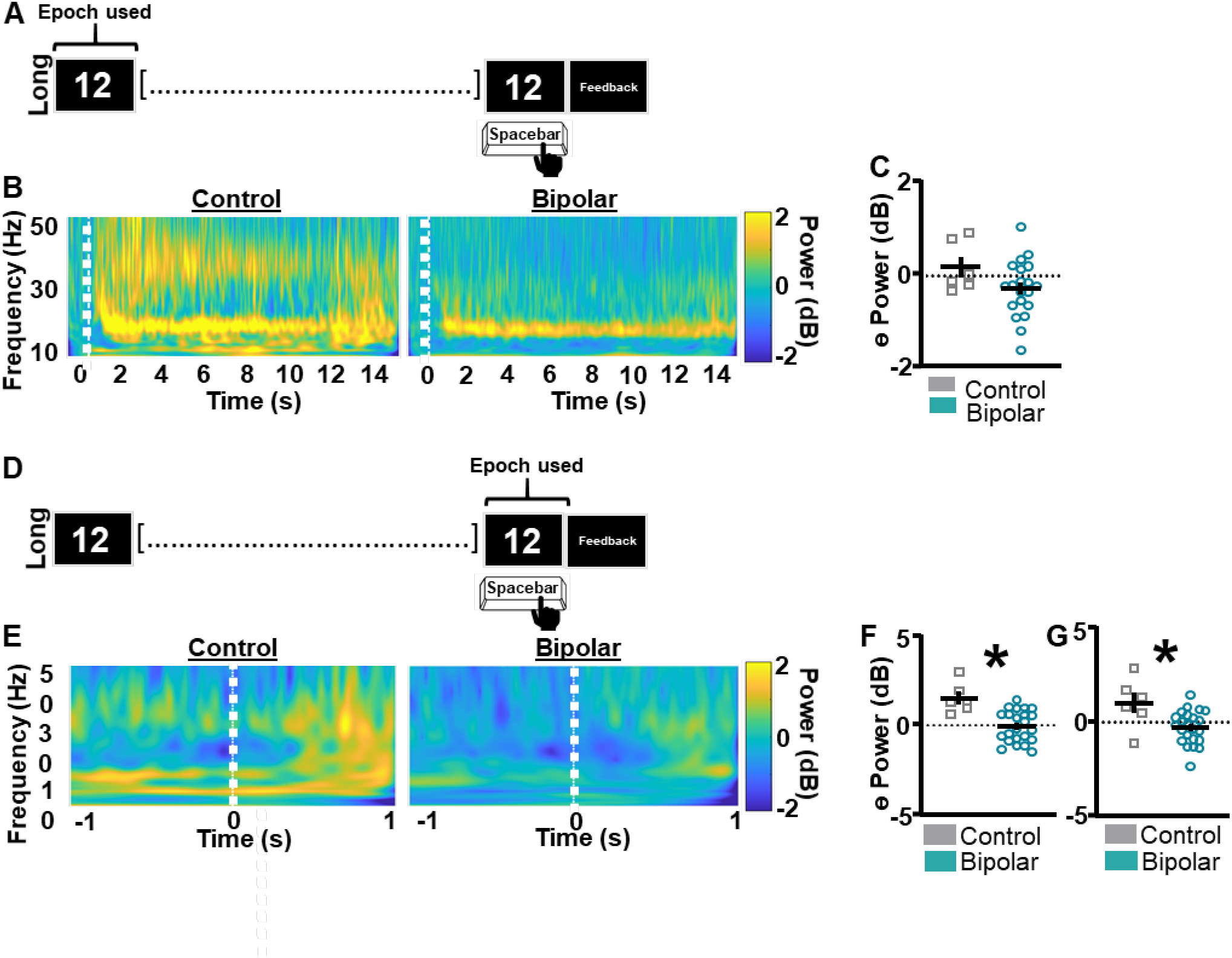
Theta power surrounding the long interval response is lower in the bipolar group compared to the control group. **A**. Data were epoched around the presentation of the long interval timing cue. **B**. Averaged spectrogram of individuals in the control group **[left]** and the bipolar group **[right]**. Exploratory analyses suggest that oscillatory activity does not differ between the two groups during the whole long interval epoch. **C**. ROI-based analyses indicate that theta power following the timing cue does not differ between bipolar and control groups. **D**. Data were epoched around the long interval button press. **E**. Averaged spectrogram of individuals in the control group **[left]** and the bipolar group **[right]**. Exploratory analyses suggest that oscillatory activity does not differ between the two groups. **F**. ROI-based analyses indicate that theta power prior to the response was lower in the bipolar group compared to the control group. **G**. ROI-based analyses indicate that theta power following the response was lower in the bipolar group compared to the control group. Mean and standard error of the mean plotted in bar graphs. Dots represent values from individual subjects. * p < 0.05

Data were next epoched surrounding the button press indicating the participants’ estimation of long interval duration (**Figure 4D**). Averaged spectrograms were created for individuals in the CT group (**Figure 4E[left])** and the BD group (**Figure 4E[right]**). Exploratory analyses suggest that oscillatory activity did not differ between groups. ROI-based analyses suggest that individuals in the BD group showed lower theta power compared to the CT group before (*t*_(27)_ = 3.946, *p* = 0.0005, one BD outlier excluded, **Figure 4F**) and after (*t*_(27)_ = 2.852, *p* = 0.0082, one BD outlier excluded, **Figures 4G**) the response.

### Timing performance and frontal theta within BD sub-groups

Because a lacuna exists in the literature regarding how mood symptoms and medication status affect cognitive symptoms associated with BD, our next set of planned analyses focused on differences in timing performance and frontal theta between sub-groups of individuals with BD.

The epoch of data analyzed included the entirety of the ITT (**Figure 5A**). Response curves suggest that supra-second ITT performance is not significantly associated with differences mood state (**Figure 5B**). Peak time and CV indices did not differ between groups for the short interval (Peak time: *t*_(20)_ = 0.05827, *p* = 0.9541; CV: *t*_(21)_ = 0.3629, *p* = 0.7203, one BD outlier excluded, **Figure 5C**) or the long interval (Peak time: *t*_(21)_ = 1.333, *p* = 0.1969; CV: *t*_(21)_ = 0.3012, *p* = 0.7662, **Figure 5D**). Frontal theta power also did not differ between groups as shown in **Figure 5E** (*t*_(20)_ = 0.1963, *p* = 0.8463, one BD outlier excluded).

**Figure 5.**
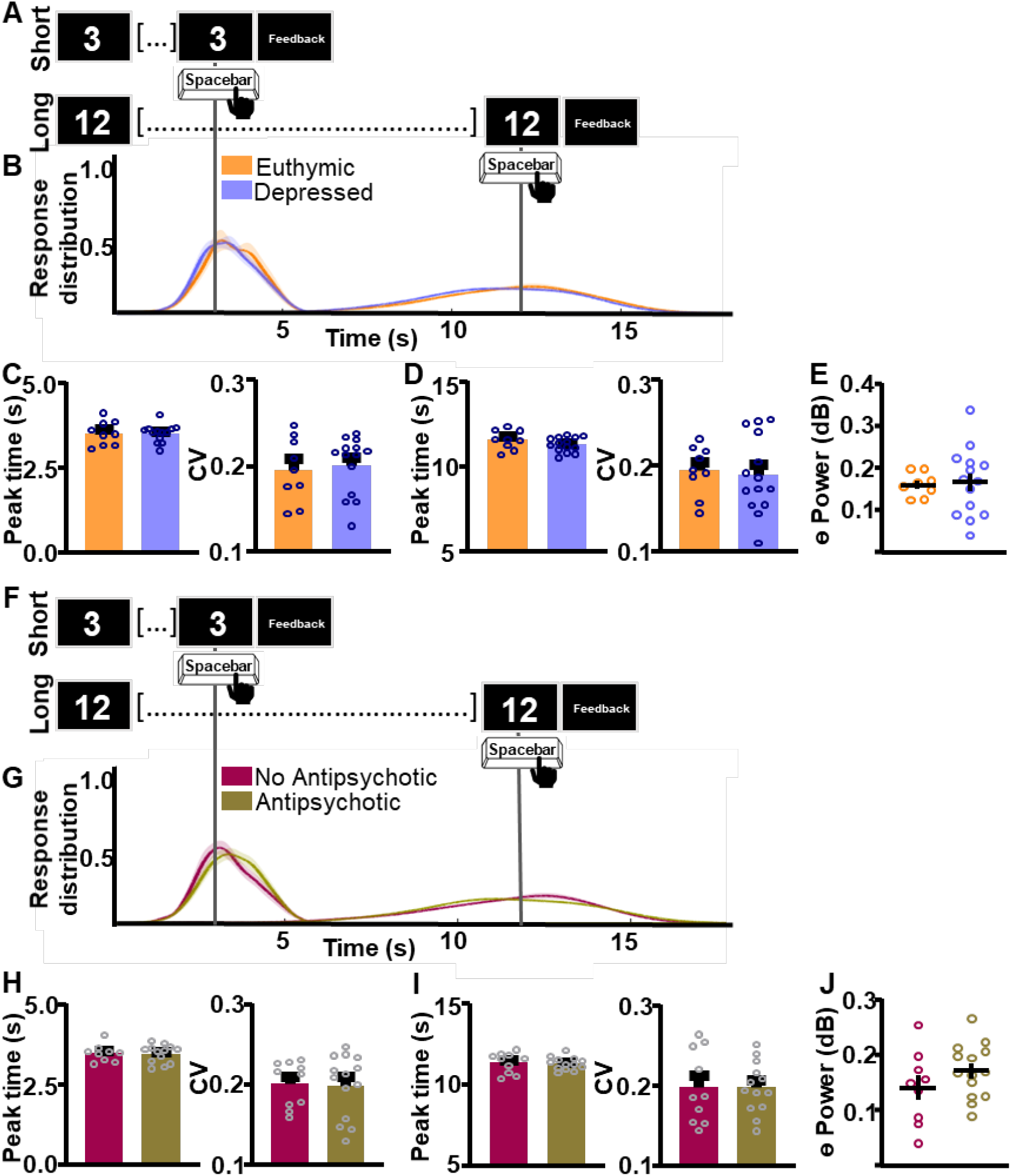
Interval timing performance and frontal theta power do not differ as a function of mood or medication status. **A**. To assess task-wide differences in oscillatory activity data from the whole interval-timing task were analyzed. **B**. Response distribution for individuals with bipolar disorder who were either euthymic or depressed at the time of data collection. **C-D**. Groups do not differ in time estimation for the short **[C]** or the long **[D]** intervals. **E**. Frontal theta power during the ITT did not differ between groups. **F**. Response distribution for individuals with bipolar disorder divided by anti-psychotic medication status. **G-H**. Groups do not differ in time estimation for the short **[G]** or the long **[H]** intervals. **I**. Frontal theta power during the ITT did not differ between groups. Mean and standard error of the mean plotted in bar graphs. Dots represent values from individual subjects.

The epoch of data analyzed included the entirety of the ITT (**Figure 5F**). Response curves suggest that supra-second ITT performance is not significantly associated with differences in antipsychotic medication status use (**Figure 5G**). Peak time and CV indices did not differ between groups for the short interval (Peak time: *t*_(20)_ = 0.1367, *p* = 0.8927; CV: *t*_(22)_ = 0.1525, *p* = 0.8802, **Figure 5H**) or the long interval (Peak time: *t*_(21)_ = 0.2647, *p* = 0.7938, one BD outlier excluded; CV: *t*_(21)_ = 0.02555, *p* = 0.9799, one BD outlier excluded, **Figure 5I**). Frontal theta power during the ITT also did not differ between groups **Figure 5J** (*t*_(21)_ = 1.284, *p* = 0.2133, one BD outlier excluded).

## Discussion

The objective of the present work was to assess supra-second ITT performance in individuals with BD. Results suggest that task performance was impaired in the BD group compared to the CT group. Additionally, frontal theta power in the BD group during the ITT, was significantly lower than that in the CT group. We also examined if mood state or medication status affected ITT performance within the BD group. Results suggest that neither ITT performance nor frontal theta were significantly different in association with these factors.

### Neuroanatomy of interval timing and what this suggests about the pathophysiology of BD

For decades, neuropsychologists have used behavioral testing to identify which brain regions/networks subserve differential functioning in the brains of patients. However, using the results of ITT performance to triangulate *single regions* which are abnormal in BD presents a challenge, as the neuroanatomy of time processing is famously diffuse^17^. Indeed, no single brain region processes the passage of time, rather, this ability comes from the coordinated functioning of a network involving multiple brain regions and neurotransmitter systems.

One possible neural mechanism underlying the altered ITT performance observed in the present work is the abnormal functioning of the dopamine system in individuals with BD. Indeed, the dopamine hypothesis of BD, which proposes intrinsic dysregulation of dopamine receptor transporter homeostasis in patients^3,31^, is widely used to explain the pathophysiology associated with the disorder. Additionally, abnormalities in supra-second interval processing have been reported in other neuropsychiatric conditions where alterations of dopaminergic pathways play an important role, including SCZ, Parkinson’s disease and Huntington’s disease^17^.

Another possible neural mechanism subserving the ITT performance and the frontal theta deficits identified in the present work are the well-characterized frontal cortical abnormalities observed in individuals with BD including reductions in cortical grey matter^18,32^. Indeed, compromised frontal cortical activity has been linked to abnormalities in supra-second interval timing^17^. Interestingly, more recent work has also identified cerebellar abnormalities in BD including altered cerebellar metabolism^19^, reduced vermal volume, reduced cerebellar grey matter^18^, abnormal cerebello-frontal connectivity^20^, and reduced neurotrophic signaling^21^. A substantial body of work suggests that the cerebellum is a key region in sub-second interval timing^33-35^. Additionally, recent work suggests that cerebellar activity is also related to supra-second interval timing^25,36^. Because of the wide-ranging impairments individuals with BD show in interval timing, from implicit motor, to sub-second to supra-second, further work should explore if modulation of cerebellar activity in BD could rescue these deficits.

### Frontal theta abnormalities during the ITT

Previous work from our group suggests that reduced theta power in the 500ms following timing-cue presentation is related to abnormal supra-second ITT performance in SCZ patients^25^. Because of the genetic and symptomatic overlap between SCZ and BD^4^, we hypothesized that individuals with BD would show similar impairments. Indeed, the present work identified abnormalities in ITT performance and in theta power in BD patients; however, oscillatory abnormalities were not time-locked to the post-cue interval as they were in SCZ. Instead, our work suggests that, while theta power increased during the performance of the ITT compared to rest for the CT group, as is expected, no such increase was seen in the BD group.

Differences in how frontal theta abnormalities present between SCZ and BD groups might be linked to differences in extent and type of cognitive impairments between the two disorders, as well as differences in the extent of neuropathological changes associated with each disorder. Relatively global cognitive impairments and diffuse brain volume reductions^37^ accompany SCZ; indeed, some researchers propose that this may be the core pathology underlying the condition^4^. Conversely, in BD, the cognitive impairments and neuropathological abnormalities are less well characterized, less severe, and may affect different domains than in SCZ^4,7^. It is thus reasonable to conclude that the neural basis of these cognitive impairments is condition-specific and produces distinct differences in the timing of oscillatory activity. Indeed, differences in EEG oscillatory activity are seen between groups of patients with SCZ and BD during other tasks and at rest^38^. Further investigation into these disorder-specific EEG abnormalities could result in interesting diagnostic and neuropathological insights.

One surprising finding from the present dataset is that individuals with BD showed lower theta power surrounding the response for the long interval, but not the short interval. Task performance data indicate that the types of impairments shown by individuals with BD differ between short and long intervals. For short interval, participants show altered accuracy, while for the long interval participants show altered precision. This is not an uncommon finding, as the long interval is more difficult to estimate. These results could indicate that frontal theta power is more closely linked with response distribution (precision) than response accuracy. However, further work is necessary at this point to substantiate this claim.

### Trait vs. state cognitive abnormalities in BD

Debate exists in the literature regarding which cognitive impairments associated with BD are state dependent, i.e. dependent on mood or medication status, and which are trait characteristics of BD. Our results indicate that supra-second timing performance is not altered as a function of mood or antipsychotic medication status, thus suggesting that deficits in supra-second interval timing might be a trait marker of BD. Together with previous work indicating that individuals with BD show impairments in sub-second^13,15^ and implicit motor timing^14^, results suggest that altered ability to assess the passage of time across all time scales is a key trait of BD.

## Data Availability

All data produced in the present study are available upon reasonable request to the corresponding author.

## Acknowledgements

The authors would like to acknowledge Laren Garrett for her excellent technical assistance.

