## Supplemental materials for "Supra-second interval timing deficits and abnormal frontal theta oscillations in individuals with bipolar disorder"

**Supplemental materials for:** Supra second interval timing deficits and abnormal frontal theta activity in individuals with bipolar disorder

Supplemental results

Response curves suggest that individuals with BDI and BDII do not differ in their supra-second ITT performance (**Supplemental Figure 1A-B**). Peak time and CV indices do not differ between groups for either the short (Peak time: $t_{(22)}$ = 0.02449, $p$ = 0.9807, **Supplemental Figure 1C [left]**; CV: $t_{(22)}$ = 0.4230, $p$ = 0.6764, Supplemental **Figure 1C [right]**) or long (Peak time: $t_{(22)}$ = 0.9181, $p$ = 0.9277, **Supplemental Figure 1D [left]**; CV: $t_{(22)}$ = 0.3181, $p$ = 0.7534, Supplemental **Figure 1D [right]**) intervals. Additionally, theta power during the ITT does not differ between individuals with BDI vs. BDII ($t_{(21)}$ = 1.268, $p$ = 0.2188, one BD outlier excluded, **Supplemental Figure 1E**).

The epoch of data analyzed included the entirety of the ITT (**Supplemental Figure 2A**). No differences in power were detected between the BD and CT groups during the ITT for the following frequency bands: delta ($t_{(28)}$ = 1.607, $p$ = 0.1193, **Supplemental Figure 2B**), alpha ($t_{(27)}$ = 0.1129, $p$ = 0.9109, one BD outlier excluded, **Supplemental Figure 2C**), beta ($t_{(28)}$ = 0.7105, $p$ = 0.4832, **Supplemental Figure 2D**), and gamma ($t_{(27)}$ = 0.6337, $p$ = 0.5316, one BD outlier excluded, **Supplemental Figure 2E**).

Supplemental Figures

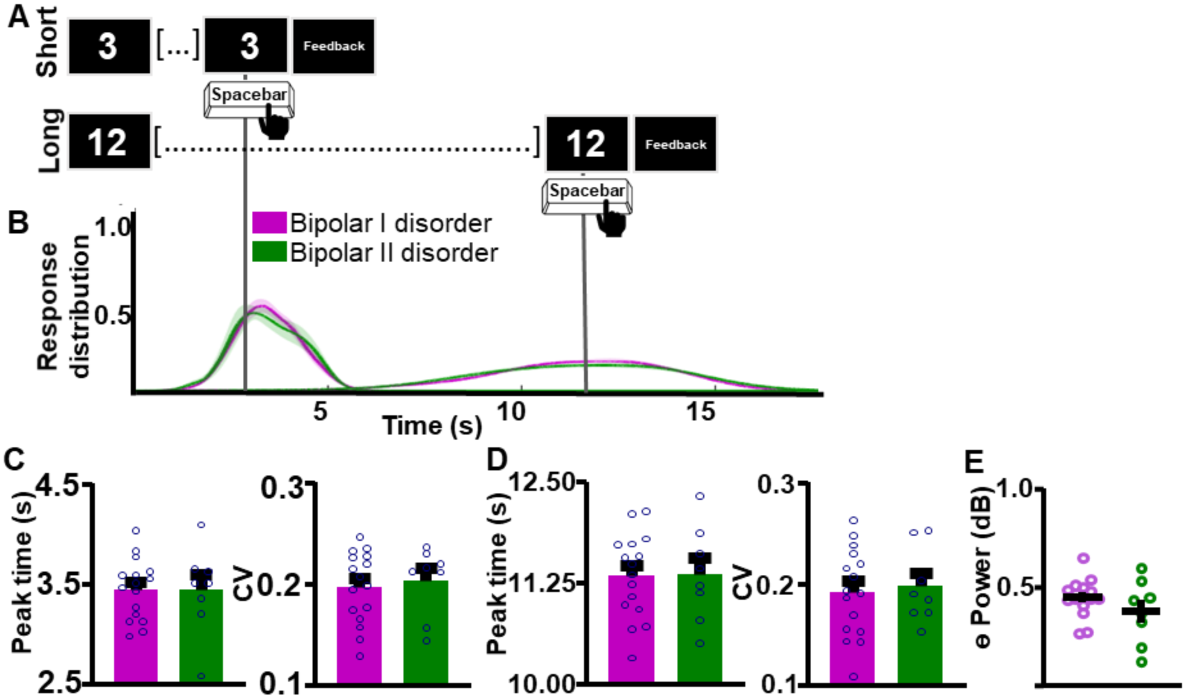


**Supplemental Figure 1. Interval timing performance and frontal theta power do not differ between individuals diagnosed with Bipolar I vs. Bipolar II disorder. A.** Schematic diagram of supra-second interval timing task. Trials begin when participants are shown a 3s or a 12s timing cue. Participants press a button to indicate their estimation of the target interval. **B.** Response distribution for individuals with bipolar I or bipolar II disorder. **C-D.** Groups do not differ in time estimation for the short **[C]** or the long **[D]** intervals. **E.** Frontal theta power during the ITT did not differ between groups. Mean and standard error of the mean plotted in bar graphs. Dots represent values from individual subjects.

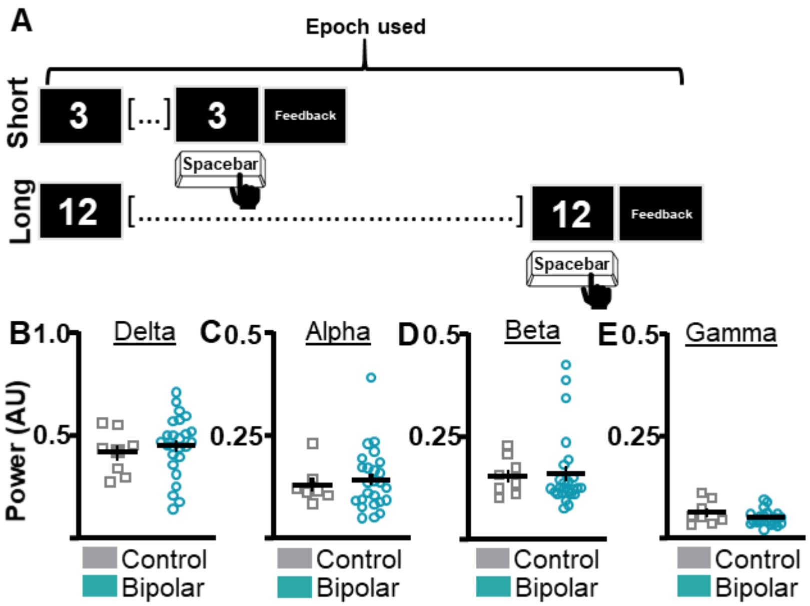


**Supplemental Figure 2. Power in frequency bands other than theta did not differ between bipolar and control groups. A.** To assess task-wide differences in oscillatory activity between bipolar disorder and neuronormative control groups data from the whole interval-timing task were analyzed. **B-E.** No differences in power were detected between bipolar and control groups for the following frequency bands: delta **[B]**, alpha **[C]**, beta **[D]**, and gamma **[E]**. Mean and standard error of the mean plotted in bar graphs. Dots represent values from individual subjects.
